# Epigenomic profiles of African American *Transthyretin* Val122Ile carriers reveals putatively dysregulated amyloid mechanisms

**DOI:** 10.1101/2020.04.15.20066621

**Authors:** Gita A Pathak, Frank R Wendt, Antonella De Lillo, Yaira Z. Nunez, Aranyak Goswami, Flavio De Angelis, Maria Fuciarelli, Henry R Kranzler, Joel Gelernter, Renato Polimanti

**Author notes:** Corresponding author: Renato Polimanti, PhD, Address: VA CT 116A2, 950 Campbell Avenue, West Haven, CT, 06516, USA.

## Abstract

The Val122Ile mutation in *Transthyretin* (*TTR*) gene causes a rare, difficult to diagnose hereditary form of cardiac amyloidosis. This mutation is most common in the United States and mainly present in people of African descent. The carriers have an increased risk of congestive heart failure and several other non-cardiac phenotypes such as carpal tunnel syndrome, peripheral edema, and arthroplasty which are top reasons for ambulatory/outpatient surgeries in the country. We conducted first-ever epigenome-wide association study in Val122Ile carriers of African descent for heart disease (HD) and multiple outpatient surgeries (OS) - an early disease indicator. Five differentially methylated sites (p≤2.1e-08) in genes – *FAM129B, SKI, WDR27, GLS*, and an intergenic site near RP11-550A5.2 and one differentially methylated region containing *KCNA6* and *GALNT3* (p=1.1e-12) were associated with HD. For OS, we observe four sites – two sites in *UBE2E3* and *SEC14L5*, and other two in intergenic regions (p≤1.8e-07) and three regions overlapping *SH3D21, EVA1B, LTB4R2* and *CIDEB* (p≤3.9e-07). Functional PPI module analysis identified *ABCA1* (p=0.001) for HS. Six cis-mQTLs were associated with one of the significant CpG sites (*FAM129B*; p=4.1e-24). We replicated two CpG sites (cg18546846 and cg06641417; p<0.05) in an external cohort of biopsy-confirmed cases of TTR amyloidosis. The genes identified are involved in transport and clearance of amyloid deposits (*GLS, ABCA1, FAM129B*); cardiac fibrosis (*SKI*); and muscle tissue regulation (*SKI, FAM129B*). These findings highlight the link between a complex amyloid circuit and diverse symptoms of Val122Ile.

## 1. Introduction

Hereditary transthyretin amyloidosis, caused by specific disease-causing mutations, is due to a gradual extracellular deposition of amyloid in multiple tissues primarily leading to several clinical signs and symptoms^1^. There are 113 known mutations in the transthyretin (*TTR*) gene^2^ giving rise to hereditary form of *TTR* amyloidosis. The tetrameric structure of TTR protein dissociates into dimers and monomers resulting in formation of fibrils. The aggregation of fibrils leads to deposition of “amyloid-fibrils” in multiple tissues causing onset of heterogenous symptoms ^2,3^. Val122Ile is the most prevalent *TTR* gene mutation in the United States and observed in populations of African descent^4^. The Val122Ile is a point mutation (variant - rs76992529) resulting in substitution of isoleucine with valine at 122 position. Extensive amyloid deposition seems to resemble hypertrophic cardiomyopathy such as enlargement or wall thickening leading to heart failure and atrial fibrillation^5^. These symptoms often are attributed to other population-prevalent cardiovascular risk factors resulting in underestimation of the clinical penetrance of the Val122Ile^6^. The estimated age of onset for non-cardiac precursor phenotypes for hereditary transthyretin amyloidosis is between 30 to 40 years of age^7^. In two retrospective studies, carpal tunnel syndrome preceded hereditary transthyretin amyloidosis diagnosis by 9-10 years^8,9^. According to The Transthyretin Amyloid Outcome Survey (THAOS), several cardiac, gait, gastrointestinal, neurological and renal disorders are prevalent in Val122Ile carriers^4^. Parallel to these findings, another study reported phenotypes associated with TTR mutations such as atrial fibrillation, myopathy related to ventricular thickness, gastrointestinal and kidney dysfunction including nausea, vomiting, and neuromuscular dysfunction^10^.

Recently, we reported a combined association of heart disease history and having had 10 or more outpatient (ambulatory) surgeries with Val122Ile mutation in individuals of African descent^11^. One of the top reasons for ambulatory surgery in the United States is arthroplasty^12^, which occurs in TTR-carriers years before the expected cardiac dystrophy at advanced ages^13^. These epidemiological findings indicate that atypical phenotypes occurring earlier in life could be connected to the risk of heart failure in Val122Ile carriers. Findings from several studies including ours raise the possibility of non-regulatory molecular factors contributing to the genotype-phenotype correlation^10,14–17^. Therefore, understanding the underlying biological changes in Val122Ile carriers is key to explaining the symptom heterogeneity and earlier onset of atypical phenotypes.

DNA methylation is a heritable non-coding regulatory mechanism causing phenotypic variation^16^. Epigenetic modifications arising from the addition of methyl groups on cytosine-phosphate-guanosine (CpG) sites^18^ could contribute to molecular mechanisms involved in *TTR* amyloidosis. So far, no study has investigated this hypothesis. Aberrant methylation profiles have been implicated in increasing accelerating the progression of common and rare diseases^19^. The accumulation of amyloid-fibrils within or around cellular structures of the tissue result in damage invoking an immune response^20^. The inter-individual variation in response to site of damage invokes an acute phase response^20^. DNA methylation profiles have the potential to capture individual-level variability and highlight mechanisms involved in *TTR* amyloidosis^21^. Thus, we conducted the first epigenome-wide association study of *TTR* Val122Ile carriers to investigate the association of methylation changes with medical history of heart disease and outpatient surgeries.

## 2. Methods

### 2.1 Cohort description

Carriers of Val122Ile (rs76992529*G>A) risk allele were selected from the Yale-Penn cohort,^22–25^ whose medical history was obtained as part of the phenotyping effort, as previously reported^11^. The Yale-Penn study was approved by the institutional review boards at each participating site. The current study was approved under the protocol 2000023750 by the institutional review board (IRB) at Yale University School of Medicine.

We investigated two binary phenotypes – self-reported history of heart disease (“*Has a doctor ever told you that you have (had) a heart disease?*”) and having had 10 or more outpatient surgeries. We previously reported that these two phenotypes were associated with the Val122Ile mutation in the Yale-Penn cohort^11^. We assayed 104 African Americans, samples from 8 of which failed the assay, leaving 96 carriers for analysis (mean age = 42.16 ± 9.9 [SD] yrs.). Epigenomic differences were tested with respect to the history of heart disease (N=90 controls; 6 cases; Males-47%) and 10 or more outpatient surgeries (N=94 controls, 2 cases; Males-46%).

The significant methylation sites were investigated in an independent cohort of Italian participants of European descent. This study included 48 carriers of *TTR* mutations (Val30Met, Phe64Leu, Ile68Leu, Ala120Ser, and Val122Ile) with the diagnosis of TTR amyloidosis confirmed via positive amyloid fibril deposition in their cardiac tissue biopsy samples, in addition to clinical symptoms. Thirty-two healthy individuals with none of the *TTR* mutations or clinical symptoms from the same local area were recruited as controls. Detailed information regarding this cohort was previously reported^14,26^. The methylation array analysis was performed at the Connecting Bio-research and Industry Center, Trieste, Italy, using the same sample protocol implemented in the Yale-Penn cohort.

### 2.2 Sample and array processing

DNA was extracted from whole blood of Yale-Penn participants using the EZ-96 DNA methylation kit (Zymo Research, CA, USA). As previously described^25^, the samples were genotyped at the Yale Center for Genome Analysis (YCGA), the Center for Inherited Disease Research, and the Gelernter laboratory at Yale (VA CT) using genome-wide arrays (Illumina HumanOmni1-Quad v1.0 and Illumina HumanCoreExome arrays), the imputation was performed with IMPUTE2.0, and principal components were derived on the QC’d genomic data. The Val122Ile – rs76992529 probe was genotyped in the HumanCoreExome array and imputed in the HumanOmni1-Quad v1.0 array with high imputation quality (INFO score = 0.98) to determine TTR carriers in the African-American Yale-Penn participants^11^. DNA methylation was assayed using the Illumina Infinium MethylationEPIC chip quantifying >850,000 CpG sites and imaged on the Illumina iScan system at YCGA. The methylation intensity data (*.idat files) was exported for analysis using the manufacturer’s recommended protocol using the GenomeStudio methylation module.

### 2.3 DNA methylation analysis

All analyses were performed in R 3.6. The methylation intensity files (*.idat) were imported into *ChAMP* ^27^ for post-processing and normalization. The beta values, ranging from 0 to 1 were generated for all CpG sites representing the ratio of methylated to unmethylated fluorescent intensities. Primary QC removed CpG sites with low detection p-value, missing beads, sites near SNPs, multi-hit and non-autosomal sites. The beta values of the remaining 737,385 sites were normalized using the beta mixture quantile (BMIQ) method, followed by analysis of batch effects using singular vector decomposition. Technical batch effects for array and slide were corrected using ComBat. The blood cell-type composition and smoking status^28^ was derived from methylation data. The association analysis for CpG sites was performed on M-values (transformed beta values) using empirical Bayes methods implemented in the *limma* package. The association was adjusted for age, sex, tobacco use, smoking, genotype-derived principal components 1-10, and blood cell type proportions: CD8+T cells, CD4+ T cells, natural killer cells, B-cells, monocytes, and neutrophils. Genomic inflation was calculated using the *QQperm* package; both phenotypic associations had a lambda of 1 or less (Heart Disease – 1.01 and Outpatient Surgery – 0.92) indicating lack of population substructure bias. Due to the unbalanced case-control proportions, we also performed permutation of the association using the *CpGassoc* package^29^. Differential methylation of regions was calculated using *DMRcate*. The sites and regions were deemed significant considering a false discovery rate of <0.05 (FDR_p-value_<0.05). Differentially expressed PPI network modules were investigated using the *FEM* package^30^. Gene ontology using significant genes mapped from significant CpG sites and regions was assessed with *ShinyGO* ^31^. The cis-methylation QTL (association between SNPs and significant methylated CpG sites) was performed with all the aforementioned covariates using *MatrixQTL*^32^, wherein the local distance was defined using the biologically expected and default value of 1 Mb between SNP and CpG sites. Significant associations were identified using genomic control adjustment (GC) p-values < 0.05, in addition to FDR_p-value_ < 0.05. Epigenetic age – defined as the cumulative score of specific CpG sites across the genome -- was calculated using Hannum and Horvath’s clock from the *wateRmelon* package^33^. As suggested by the developers of each of the epigenetic clocks, biological age, and chronological age were significantly correlated, R≥0.90; p<0.001. The difference in the epigenetic (biological) age between cases and controls was tested using Student’s t-test. The network connecting all of the significant genes detected from the above analyses was constructed using GeneMANIA^34^. The results were visualized using ggplot2, coMET^35^, and FUMA^36^.

## 3. Results

### 3.1 Differentially methylated sites

We investigated differentially methylated sites with respect to two binary outcomes: – a) self-reported heart disease and b) a history of 10 or more outpatient surgeries. After performing the recommended quality control procedure, we investigated 737,385 sites in 96 individuals. In addition to performing standard association analysis, we permuted the phenotypes (p_perm_: p-value from permutation), which accounted for the case-control imbalance, yielding nine significant CpG sites (Figure 1). Five sites were hypomethylated in individuals with heart disease: cg06641417 (*FAM129B*; logFC=-1.822; p_perm_=1.6e-08), cg26033908 (*SKI*; logFC=-1.615; p_perm_=1.7e-08), cg14890866 (*WDR27;* logFC=-2.028; p_perm_=3.0e-08), cg15522719 (*GLS;* logFC=-1.731; p_perm_=4.7e-08) and cg18546846 (intergenic; logFC=-0.786; p_perm_=2.2e-08). The CpG sites mapped to *FAM129B* and *SKI* are located in gene bodies, cg14890866 is between the 5’UTR (Un-Translated Region) and TSS200 (− 200 nt upstream of Transcription start site^37^) of *WDR27*, while cg15522719 is in TSS150 at *GLS*. Four methylation sites were associated with 10 or more outpatient surgeries: cg13998023 (*UBE2E3*; logFC=-2.632; p_perm_=1.8e-07), cg05189127 (intergenic; logFC=1.885; p_perm_=1.4e-07), cg03718655 (*SEC14L5*; logFC=-2.673; p_perm_=1.5e-07) and cg25814327 (intergenic; logFC=-2.075; p_perm_=3e-08). Three sites were hypomethylated, while cg05189127 (intergenic) was hypermethylated. The two sites that were annotated to genes were in the 5’UTR (*UBE2E3*) and TSS200 (*SEC14L5*). Details of the association result and annotation are reported in Supplementary file1 (Table S1).

**Figure 1:**
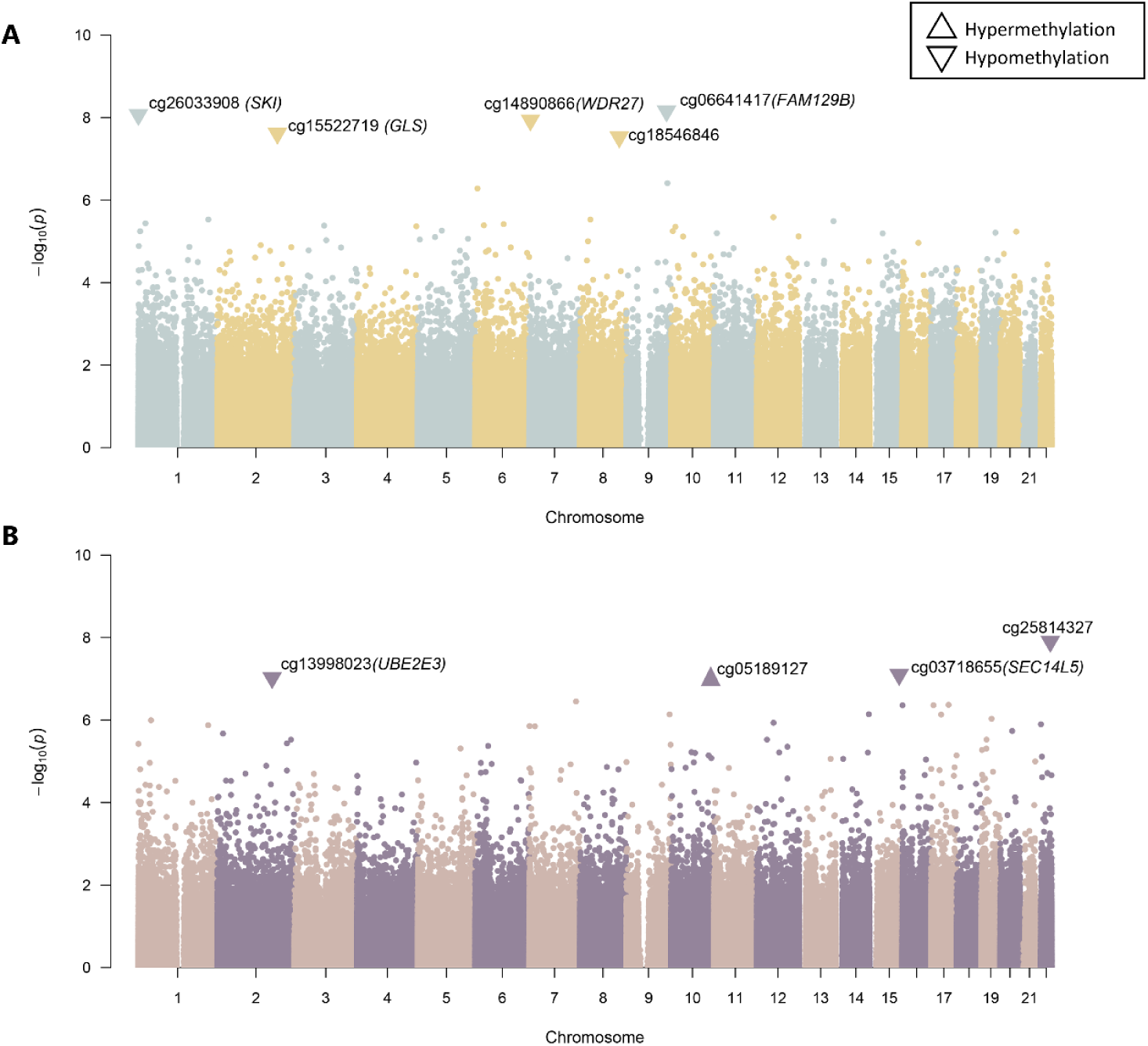
Differentially methylated sites in African American TTR-Val122Ile carriers. **(A)** Methylation sites that were significantly associated with medical history of heart disease. **(B)** Methylation sites that were significantly associated for having had 10 or more outpatient surgeries. Each CpG site is represented as a data point, with the x-axis being the genomic location and the y-axis is the -log10 of the p-value of the CpG site. Significant sites are shown as triangles and labelled with genic annotation in parentheses, triangles pointing upwards signify hypermethylation, whereas triangles pointing downwards signify hypomethylation.

### 3.2 Differentially methylated regions

For heart disease, one region on chromosome 12 overlapping *KCNA6* and *GALNT3* (p=1.1e-12) was differentially methylated. Associations with more than 10 outpatient surgeries were identified on chromosome 1 (*SH3D21; EVA1B;* p=1.3e-09), chromosome 10 (intergenic region; p=1.7e-08) and chromosome 14 (*LTB4R2; CIDEB*; p=3.9e-07). Methylation levels among all sites were positively correlated within each region. (Figure 2; Supplementary file 1; Table S2)

**Figure 2:**
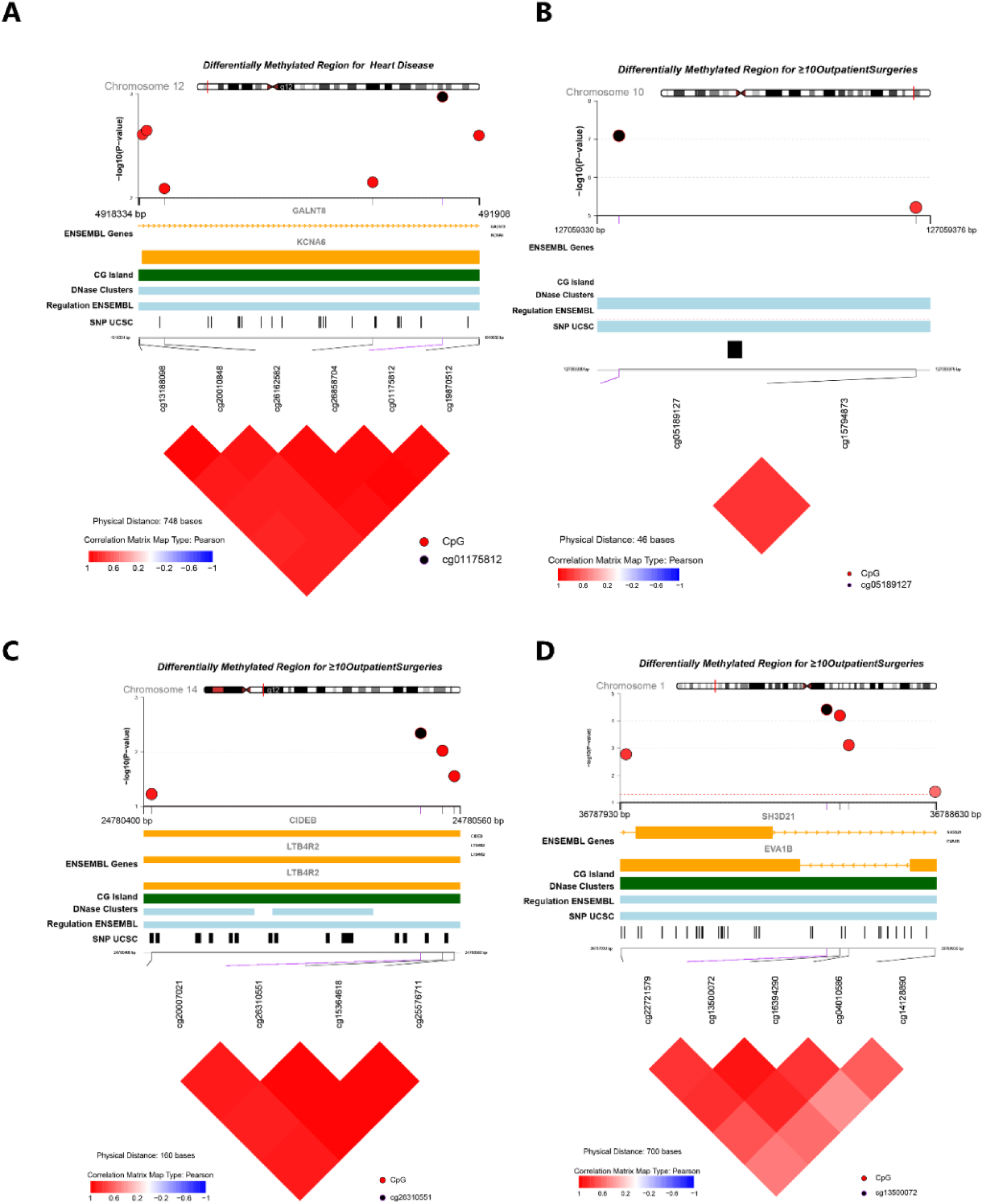
Differentially methylated regions in TTR-Val122Ile carriers. **(A)** Regional association with heart disease **(B-D)** Regional association with 10 or more outpatient surgeries. Each panel displays the association of sites within each region, followed by genomic location, ENSEMBL gene name, DNAse, regulation, SNP tracks (from UCSC browser) and correlation of CpG sites shown as a heatmap.

### 3.3 Overrepresented gene ontology and PPI networks

Differentially methylated sites and regions were annotated to their respective genes using UCSC RefGene: hg19 genome build. The gene ontology (GO) analysis identified 15 significantly enriched pathways in GO’s biological process. *GLS, SKI, GALNT8*, and *KCNA6* are involved in protein oligomerization (FDR_p-value_=4.8e-03) and *KCNA6* and *GALNT8*, which are located near one another, are involved in potassium ion transport (FDR_p-value_=4.9e-02). *FAM12B* and *SKI* (FDR_p-value_=4.8e-03 to 3.2e-02) participate in the development of various tissue types – myotubules, and skeletal and striated muscles (Figure 3).

**Figure 3:**
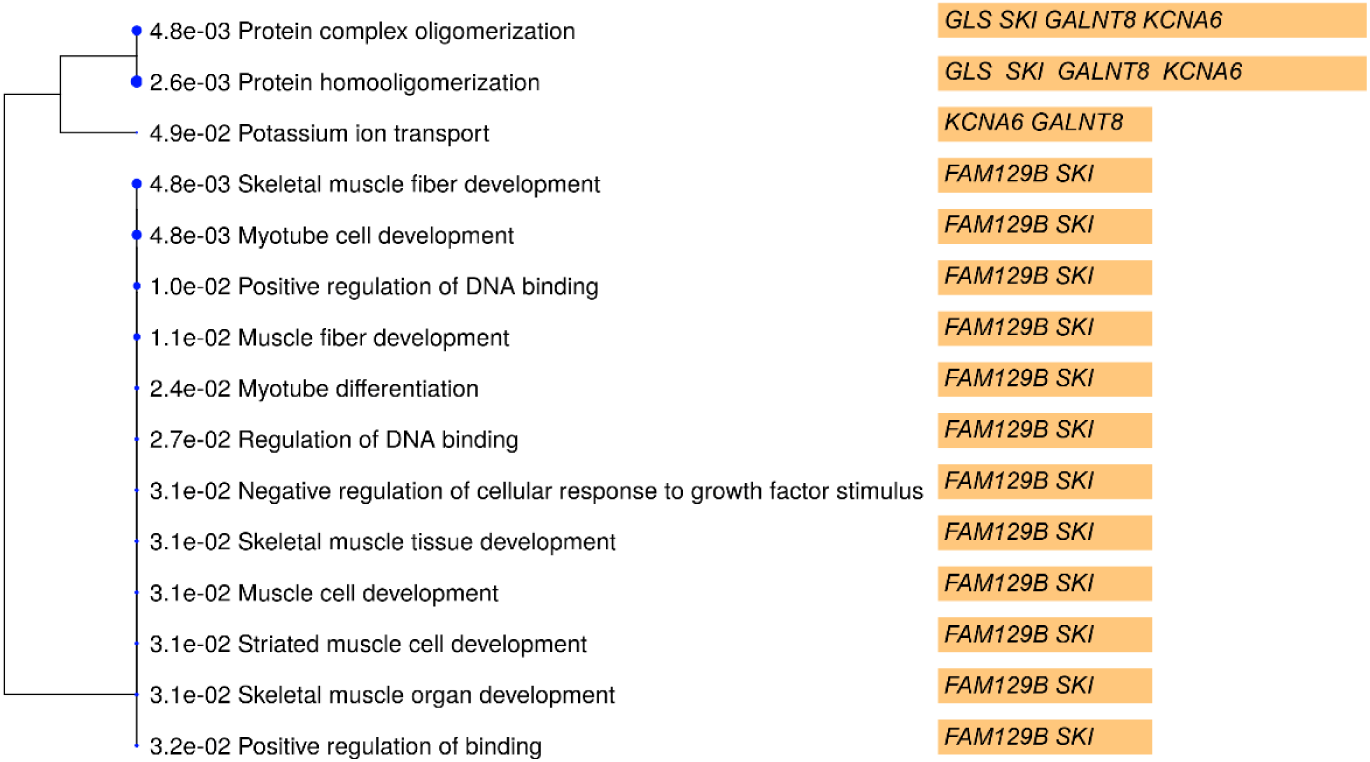
Enriched gene ontology (GO) biological processes. The dendrogram shows the FDR_pvalue_ of the pathway associations and are grouped by similarity of function. The genes involved in each of the processes are highlighted in orange bars.

We also investigated the methylation sites for differentially methylated functional modules using the R package *FEM* ^*30*^. The CpG sites are weighted based on their location in the genes, which are then mapped to a protein-protein interaction (PPI) network. For each module (i.e. PPI network) identified, the seed gene is the primary gene to which other functionally related genes are connected. For heart disease, we found the *ABCA1* module to be significant (p=0.001) and target genes identified within the module were: *ABCA1, SNTB2, BLOC1S2* and *LIN7B* (p<0.05). The *EXOSC4* gene module was associated with the phenotype of 10 or more outpatient surgeries, and it was the only gene that was a target (Figure 4; Supplementary file1; Table S4).

**Figure 4:**
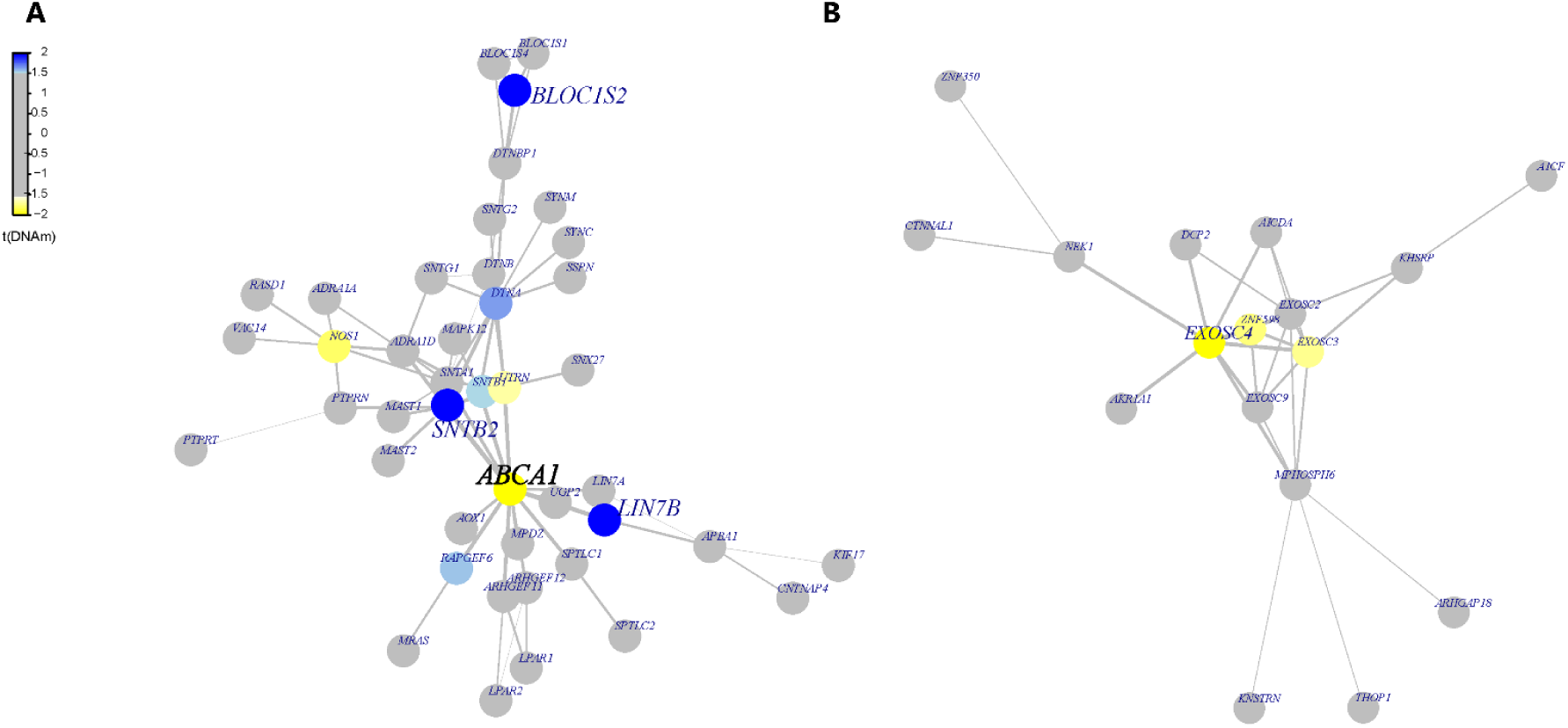
Functional Protein-Protein Interaction (PPI) Networks. The differentially methylated modules consist of a network of genes based on their functional connectivity using protein-protein interaction. Each module has primary gene which is connected to other target genes in the network. Each module was significant p<0.05 using the FEM method (see methods). The genes in blue show hypermethylation and yellow represents hypomethylation. A) ABCA1 module was associated with heart disease and the significant target genes in addition to ABCA1 were SNTB2, BLOC1S2 and LIN7B. B) EXOSC4 module was associated with outpatient surgeries and also was the only significant gene in the network.

**Figure 5:**
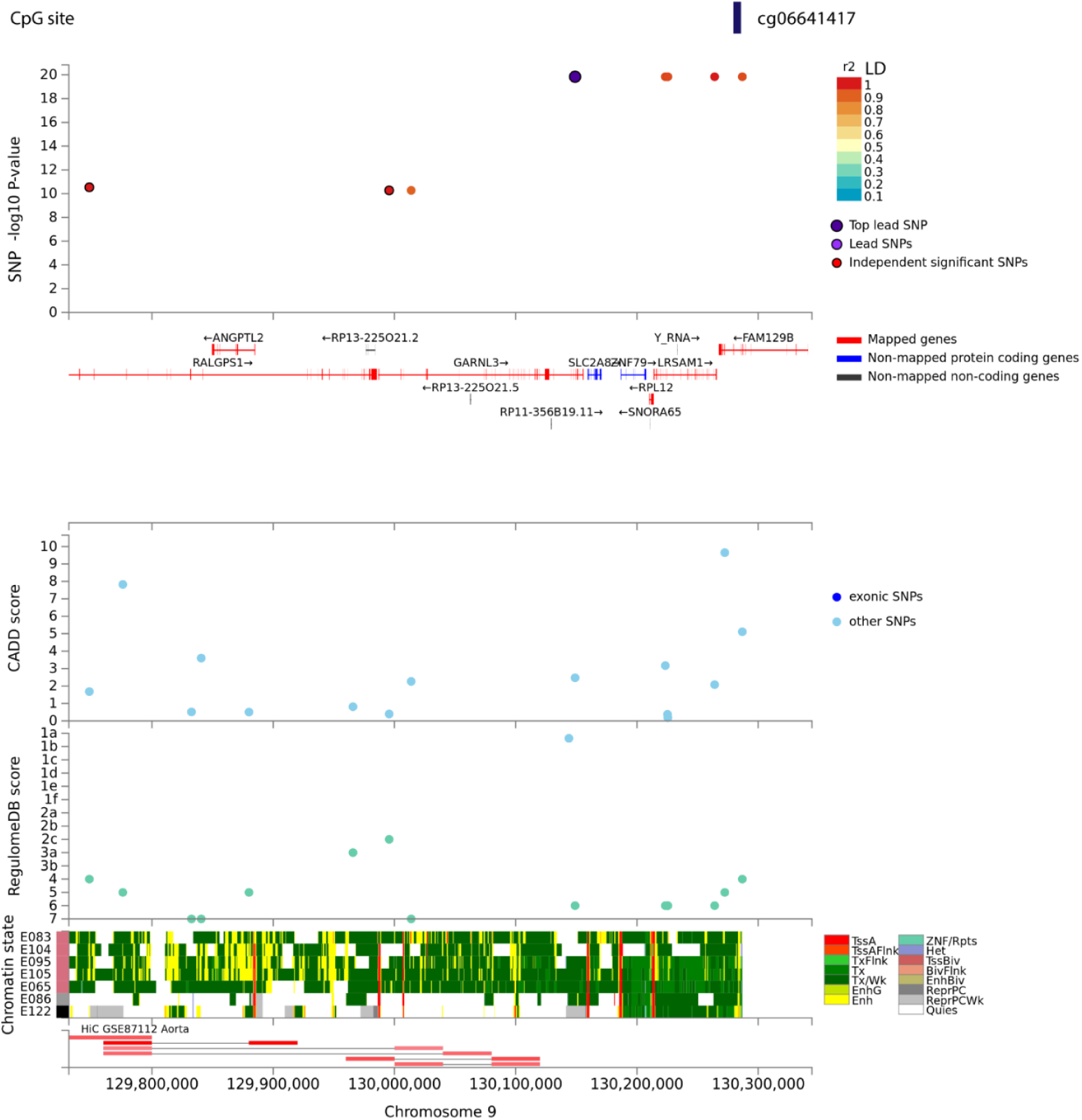
Local-mQTL associated with site – cg06641417 mapped to the FAM129B gene. The top panel displays single nucleotide polymorphisms (SNPs) associated with CpG site – cg06641417 as a data point and color coded based on linkage disequilibrium with the top lead SNP in purple. The x-axis shows gene annotation (hg19) of the region and the y-axis displays the -log10 of p-value. The following panels present various annotations of the reported SNPs i.e. CADD - Combined Annotation Dependent Depletion, and RegulomeDB – score to identify regulatory variants. The bottom panel highlights the chromatin states of various regulatory features being putatively affected from chromatin markers observed in aorta tissue cell line. Visualization made in FUMA.

**Figure 6:**
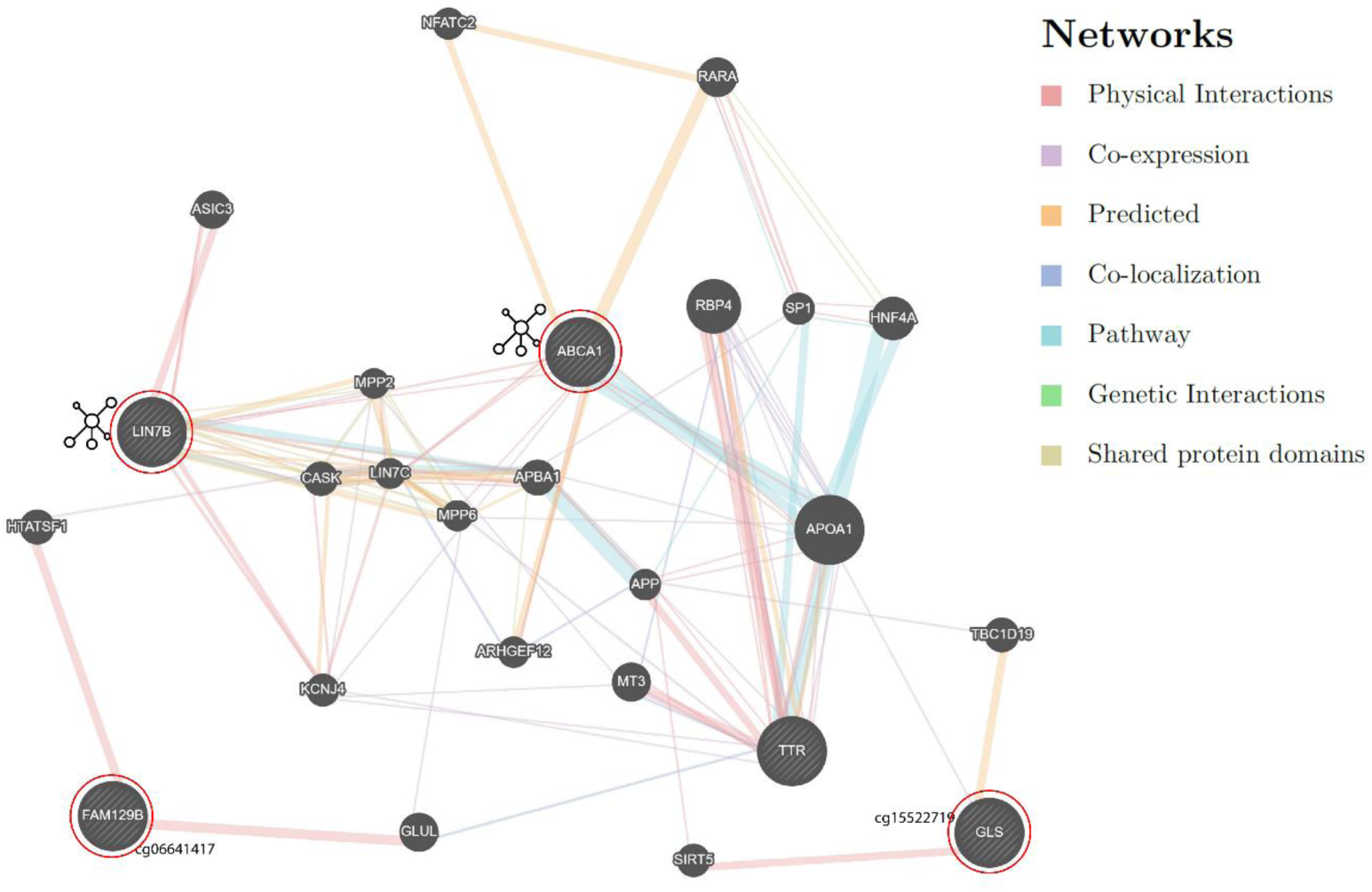
Network of genes for significant genic hits and their relationship with TTR. The genes (query genes) circled in red were identified from our analysis. The network shows intermediate genes that connect our query genes. Detailed information on the is provided in Supplementary file2

### 3.4 Local quantitative trait loci for methylated sites (mQTL)

We tested SNP associations with nine methylation sites that were epigenome-wide significant with the two phenotypes investigated. The cis-mQTL loci were defined as SNPs within ±1 Mb of the significant CpG site. The sites were considered significant based on an FDR_p-value_ < 0.05 and genomic corrected p-value (p_gc_<0.05). We found six SNPs, rs192528579, rs182192023, rs114553373, rs187644239, rs114896522, and rs139996037 significantly associated (p=4.1e-24) with site cg06641417. The SNPs are in high linkage disequilibrium (LD>0.8), rs192528579 is in the intronic region of neighboring gene – *GARNL3*; rs182192023, rs114553373, rs187644239 and rs114896522 map to *LRSAM1*. Rs139996037 is a non-coding transcript variant of the *FAM129B*.

### 3.5 Epigenetic age

The epigenetic age (DNAm) was measured using the biological clock developed by Horvath and colleagues, which uses 353 CpG sites^38^ and also with a second clock based on 71 CpG sites from Hannum and colleagues^39^. The ‘Horvath’ clock is considered to be a pan-tissue epigenetic clock, while the ‘Hannum’ clock is considered to be accurate for whole-blood tissue^40^. Both clocks estimated that carriers with heart disease are of older epigenetic age than carriers without heart disease (p_Horvath_=0.007 and p_Hannum_=0.0009) (Supplementary file1; Table S6).

### 3.6 Replication of methylation sites in the Italian cohort

We tested the nine CpG sites identified in Val122Ile carriers in an independent cohort of biopsy-confirmed *TTR* amyloidosis cases and healthy controls. We replicated cg18546846 (intergenic; near to RP11-550A5.2; p= 0.021) and cg06641417 (*FAM129B*; p=0.016) at nominal significance (p<0.05).

## 4. Discussion

The clinical consequences of the *TTR* Val122Ile mutation remain underappreciated and the syndrome that accompanies this risk mutation, under-diagnosed. Individuals exhibiting early *TTR*-amyloidosis symptoms are more likely to be diagnosed with another condition prior to receiving the diagnosis of *TTR*-amyloidosis^41^. There is nonetheless a greater burden over time towards developing ventricular hypertrophy, reduced left ventricular ejection fraction, and atrial dilation, at a later age^3,5^. We previously showed that African-American carriers of the Val122Ile mutation had a higher prevalence of heart disease and having multiple outpatient surgeries than individuals without the mutation^11^. In the present study, we identified methylation changes associated with these same phenotypes in Val122Ile carriers. We also replicated two of our CpG sites near RP11-550A5.2 and in *FAM129B* at nominal significance in a cohort that included confirmed cases of *TTR* amyloidosis. Thus, we hypothesize that the epigenetic changes associated with the pathogenesis heart disease differs from the methylation profile of carriers who are not affected by the disease. Lastly, we used GeneMANIA^34^ to interpret the interaction among the significant genes (Supplementary file2). We observed that major genes identified in the present study physically interact and share pathways with *TTR*.

*ABCA1* (ATP binding cassette transporter A1) identified via the functional network analysis encodes a transporter of cholesterol from apolipoproteins^42^. *ABCA1* regulates Apolipoprotein E (ApoE) levels, with lower expression of *ABCA1* reducing ApoE levels. However, ApoE with ApoA1 (Apolipoprotein A) reduces amyloid deposition twice as fast as inhibition of the expression of *ApoE*. Additionally, amyloid-beta levels were the lowest for the dual-knockout of *APP* (which encodes amyloid precursor protein) and *ABCA1*^43^.

*GLS* (glutaminase) is a key contributor to the metabolizing of glutamine to glutamate^44^. Amyloid-beta-treated neurons show elevated glutaminase expression, which increases glutamate levels and disrupts calcium neural regulation^45^. Additionally, neurofibrillary tangles have been shown to coexist with higher glutaminase activity^46^. The hypomethylated site in the transcription start site of the *GLS* gene may indicate its potential involvement in the central nervous system, which supports the recent finding of cerebral amyloid angiopathy in individuals with mutated TTR cardiac amyloidosis^47^. *FAM129B* (aliases; *MEG-3* and *NIBAN2*) is downregulated in tissues with amyloid deposition and animal studies have shown that enhancing the expression of *FAM129B* reduces oxidative damage by reducing amyloid-beta production via PI3K/Akt signaling^48^. Cardiac hypertrophy increases the risk of heart failure. *FAM129B* is overexpressed in heart failure samples, and rodent experiments indicate a potential role of the gene in the apoptosis of cardiac myocytes after myocardial infarction^49^. Rescuing the expression levels of *FAM129B* reverses hypertrophic responses, thus the hypomethylation of the CpG site in *FAM129B* observed in our finding supports the overexpression of the gene in heart failure^50^. The African-American population has a high prevalence of diabetes^51^. *FAM129B* is also overexpressed in cardiomyocytes under high glucose concentration reflecting its role in diabetic cardiomyopathy^52^. Although *SKI* is an inhibitor of TGF-beta-induced fibrosis and is under expressed in cardiac fibrosis, other epigenetic modulators such as miRNAs-34a and 93-c affect both *SKI* and *TGF-beta*, targeting the inhibitory factors of *SKI*, which could rescue cardiac fibrosis^53^. The gene enrichment analysis identified a role for *FAM129B* and *SKI* in the development of myotube cells and skeletal muscle fiber and organ, and striated muscle cell development.

One of the clinical findings associated with cardiac amyloidosis is increased left ventricular wall thickness, which can lead to heart failure^54^. Electrical perturbations resulting from lower potassium repolarizing current leads to a prolonged action potential in heart failure^55^. The role of *KCNA6* and *GALNT8* is associated with potassium ion transport and the transmembrane transport complexes. One of the cardiovascular symptoms of the *TTR* amyloidosis is pronounced diastolic hypertension^6^, and diastolic dysfunction is one of the symptoms associated with transthyretin amyloidosis^56^. *WDR27* was reported to be differentially methylated in individuals with significant differences in diastolic blood pressure^26^.

Aging is a common denominator to the symptomology of Val122Ile and DNA methylation^57^. Age-related methylation changes measured via “epigenetic clocks” help to identify molecular aging and its disconnect with chronological age. The Horvath clock based on 353 CpG sites and the Hannum clock based on 71 CpG sites have been extensively replicated in various tissues^58^. While these clocks were developed using blood tissues^38,39^, Horvath’s clock is validated across multiple tissues, while Hannum’s clock is more consistent in samples originating from blood tissues. Higher epigenetically derived age has been associated with several cardiovascular disease traits. Hypermethylation of genes that are protective against heart disease, lead to cardiovascular aging and increased risk for coronary disease^59^. The dysregulation of the *ABCA1* gene, the product of which is involved in the transport of cholesterol from the periphery to liver tissue^60^ has been associated with different cardiovascular pathologies. The hypermethylation of the *ABCA1* promoter region silences its expression and is associated with coronary artery disease^61^. In contrast, the increased expression of *ABCA1* regulated by ApoA1 leads to reverse cholesterol efflux in hepatic tissue ^62^. Elevated high density lipoprotein (HDL) in the liver is a target site for serum amyloid A, an acute phase response protein that is expressed during amyloidosis^63^. The observed hypomethylation of *ABCA1* and putative increase in gene expression underscores its likely involvement in shifting the methylation milieu and could perhaps explain the cardiac symptomology in a comparatively younger group of Val122Ile carriers.

These findings provide unique insights into epigenomic contrasts related to symptomology in Val122Ile carriers. However, our study has limitations. First, we investigated the Val122Ile polymorphism only for heart disease, though it is possible that we could identify additional differences with individuals who are non-Val122Ile carriers or who present with wild-type transthyretin amyloidosis. Additionally, due to the low frequency of the disease-causing mutation investigated, our study suffers from an imbalance in the ratio of cases to controls. Although the permutation analysis accounting for this imbalance confirmed our results and we replicated two associations in an independent cohort (with mostly different risk variants), our findings would benefit from replication in a larger, more balanced study.

## 5. Conclusion

Our study is the first to explore the epigenetic changes in *TTR* Val122Ile carriers. Certain Val122Ile carriers in our study presented with heart disease earlier than usually reported by individuals affected by cardiac amyloidosis. This could be due to modifier effects accelerating the pathogenicity of Val122Ile mutation. Due to the underestimated clinical penetrance of the mutation in the African American population, we leveraged an external secondary dataset with confirmed clinical phenotype as a random population sample. The purpose of this study was to understand possible non-coding mechanisms that may explain the heterogeneous phenotypes observed in Val122Ile carriers with history of heart disease. The epigenetic changes identified affect the regulation of genes involved in the transport of amyloid and regulating striated and smooth muscle, which form key components of amyloidosis and cardiac tissue susceptibility. These findings provide higher resolution on mechanisms underlying the TTR-Val122Ile mutation.

## Data Availability

Full summary statistics of the epigenome-wide analysis will be made available post-publication by contacting the corresponding author – R.P. listed on the manuscript.

## 6. Acknowledgments

The study was supported by ‘Global ASPIRE TTR Amyloidosis Competitive Grant’ from Pfizer Inc. We are grateful to the participants of the Yale-Penn cohort, which was funded under grants RC2 <u>DA028909,</u> R01 DA12690, R01 DA12849, R01 DA18432, R01AA11330, and R01 AA017535. The investigation conducted in the Italian cohort was supported by an Investigator-Initiated Research from Pfizer Inc. to the University of Rome “Tor Vergata”. The content reported in the manuscript is solely the responsibility of the authors and does not represent the official views of the NIH or Pfizer. The funding agencies had no role in the study design, data analysis, and results interpretation of the present study.

## 7. Author contributions

R.P. and G.A.P. conceptualized the research design and R.P. received funding for the study. J.G. and H. K designed the Yale-Penn study, and H.K. and J.G. oversaw the recruitment and assessment of the Yale-Penn sample. The analysis was performed by G.A.P. All authors were involved in data interpretation, manuscript preparation, review, and critical feedback. The funding for Yale-Penn was obtained by J.G and H.K.

